# Autofluorescence Virtual Staining System for H&E Histology and Multiplex Immunofluorescence Applied to Immuno-Oncology Biomarkers in Lung Cancer

**DOI:** 10.1101/2024.06.12.24308841

**Authors:** Jessica Loo, Marc Robbins, Carson McNeil, Tadayuki Yoshitake, Charles Santori, Chuanhe (Jay) Shan, Saurabh Vyawahare, Hardik Patel, Tzu Chien Wang, Robert Findlater, David F. Steiner, Sudha Rao, Michael Gutierrez, Yang Wang, Adrian C. Sanchez, Raymund Yin, Vanessa Velez, Julia S. Sigman, Patricia Coutinho de Souza, Hareesh Chandrupatla, Liam Scott, Shamira S. Weaver, Chung-Wein Lee, Ehud Rivlin, Roman Goldenberg, Suzana S. Couto, Peter Cimermancic, Pok Fai Wong

## Abstract

Virtual staining for digital pathology has great potential to enable spatial biology research, improve efficiency and reliability in the clinical workflow, as well as conserve tissue samples in a non-destructive manner. In this study, we demonstrate the feasibility of generating virtual stains for hematoxylin and eosin (H&E) and a multiplex immunofluorescence (mIF) immuno-oncology panel (DAPI, PanCK, PD-L1, CD3, CD8) from autofluorescence images of unstained non-small cell lung cancer tissue by combining high-throughput hyperspectral fluorescence microscopy and machine learning. Using domain-specific computational methods, we evaluated the accuracy of virtual H&E for histologic subtyping and virtual mIF for cell segmentation-based measurements, including clinically-relevant measurements such as tumor area, T cell density, and PD-L1 expression (tumor proportion score and combined positive score). The virtual stains reproduce key morphologic features and protein biomarker expressions at both tissue and cell levels compared to real stains, enable the identification of key immune phenotypes important for immuno-oncology, and show moderate to good performance across various evaluation metrics. This study extends our previous work on virtual staining from autofluorescence in liver disease and prostate cancer, further demonstrating the generalizability of this deep learning technique to a different disease (lung cancer) and stain modality (mIF).

**Significance:** We extend the capabilities of virtual staining from autofluorescence to a different disease and stain modality. Our work includes newly developed virtual stains for H&E and a multiplex immunofluorescence panel (DAPI, PanCK, PD-L1, CD3, CD8) for non-small cell lung cancer, which reproduce the key features of real stains.

## Introduction

Lung cancer is one of the most frequently diagnosed cancers and a leading cause of mortality worldwide. Approximately 85% of lung cancer patients have non-small cell lung cancer (NSCLC), of which the most common subtypes are lung adenocarcinoma (LUAD) and lung squamous cell carcinoma (LUSC) [1,2]. Routine diagnosis of NSCLC requires hematoxylin and eosin (H&E) staining of tissue sections, and treatment decisions are often based on the assessment of various biomarkers. Protein biomarkers on tissue sections can be visualized by immunostaining techniques. Chromogenic immunohistochemistry (IHC) is routinely used in clinical practice [3]. Multiplex immunofluorescence (mIF) is well-suited for the precise quantification and colocalization of multiple biomarkers and is widely used in research settings [3,4].

Biomarkers characterizing the tumor and immune microenvironment are valuable for diagnosis, prognosis, and treatment decisions in NSCLC. Key biomarkers of interest include pancytokeratin (PanCK), cluster of differentiation 3 (CD3), cluster of differentiation 8 (CD8), and programmed cell death ligand 1 (PD-L1). PanCK is expressed in epithelial cells and is useful for the identification of epithelial tumors [5]. CD3 is expressed in T cells, which play an important role in the adaptive immune response. CD8 is predominantly expressed in cytotoxic T cells, but can also be found on natural killer cells and dendritic cells. PD-L1, which may be expressed in both immune cells and tumor cells, suppresses the activity of T cells by binding to the regulatory receptor programmed cell death 1 (PD-1) and contributes to the immune evasion of tumors [6].

In recent years, immune checkpoint inhibitors that block PD-1 or PD-L1 have emerged as one of the most successful treatment strategies for NSCLC. PD-L1 expression is a widely used predictive biomarker of anti-PD-1/PD-L1 immunotherapy response for NSCLC. Pathologists determine the tumor proportion score (TPS) by evaluating PD-L1 expression in tumor cells [7–10], whereas the combined positive score (CPS) considers PD-L1 expression in both immune cells and tumor cells [7,11]. The immune cell topography has also been increasingly recognized as an important predictor of immunotherapy response and disease progression, and various methods to characterize immune cell phenotypes have been proposed for various cancers [12,13]. For example, the Immunoscore classification of colorectal cancer provides a scoring system based on CD3 and CD8 cell densities [14,15].

The number of stains and molecular assays that can be performed is often limited by the availability of tissue. Variability of several factors, such as tissue preparation and staining differences between laboratories, also makes consistent inter-site analysis of IHC or IF results difficult [3,4,16]. Virtual staining is a technique that addresses these problems using computer vision to generate stained images from unstained, or differently stained, tissue. One advantage of virtual staining from unstained tissue is the non-destructive process, thereby enabling the conservation of tissue samples for other uses such as standard histochemical staining or sequencing. Several methods for imaging unstained tissue have been explored, including hyperspectral autofluorescence (AF), by which endogenous fluorophores are imaged at high spatial resolution [17,18]. Virtual stains generated from AF images have demonstrated impressive qualitative and quantitative performance in several diseases and stains [19,20]. Virtual IHC has recently been shown to be promising for HER2 in breast cancer [21] and PIN-4 in prostate cancer [22]. However, there remains a lack of virtual staining applications from unstained tissue for mIF, with recent works only demonstrating stain transfer capabilities from H&E and IHC [23,24].

In this study, we demonstrate the feasibility of generating virtual stains for H&E and mIF from AF in NSCLC, extending the application of virtual staining of unstained tissue specimens to more diseases and stains.

## Materials and Methods

### Dataset

#### Data Collection and Preparation

Formalin-fixed paraffin-embedded (FFPE) tissue blocks from 448 participants with NSCLC approved for research use were procured from two biosample vendors (Avaden Biosciences, Inc., Seattle, WA, USA; Capital Biosciences, Inc., Gaithersburg, MD, USA). Institutional review board exemption was obtained for this research. Participants were 224 male (50%) and 224 female (50%). The age of participants ranged from 37 to 89 years, with a mean age of 69 years. The diagnosis consisted of 362 LUAD (80.8%), 79 LUSC (17.6%), 4 adenosquamous carcinoma (0.9%), and 3 undetermined (0.7%) NSCLC subtypes.

Two slides from serial sections were prepared from each tissue block. Both slides were scanned with a custom-built hyperspectral fluorescence microscope which has been previously described [22], to obtain unstained AF images consisting of 20 channels. The first section was then stained with H&E and scanned with the Aperio AT2 Scanner (Leica Biosystems GmBH, Nussloch, Germany). The second section was stained with a custom mIF panel consisting of DAPI to visualize nuclei, and primary antibodies paired with Opal fluorophores targeting PanCK (AE1/AE3, Opal 520), PD-L1 (E1L3N, Opal 620), CD3 (LN10, Opal 570), and CD8 (SP239, Opal 480) (Akoya Biosciences, Inc., Marlborough, MA, USA). The stained section was then scanned with the Vectra Polaris Imaging System (Akoya Biosciences, Inc., Marlborough, MA, USA) and the component channels were spectrally unmixed with the inForm Tissue Analysis Software (Akoya Biosciences, Inc., Marlborough, MA, USA). During spectral unmixing, the residual AF signal was retained as an additional channel.

Four additional slides were also prepared from each tissue block for use as control (two slides) and reserve (two slides) slides, if needed. A subset of control slides was stained for H&E and mIF to validate the staining protocols and examine sources of contamination that were occasionally observed during scanning. Reserve slides were kept unstained, for use only in case tissue was damaged during the handling process.

#### Data Alignment and Quality Control

Due to the imaging by different systems, image pairs of the AF and corresponding stains on the same slide were globally aligned with an affine transformation using an iterative approach which has been previously described [19,22]. Image pairs that could not be successfully aligned, due to tissue distortion from the staining process, for example, were excluded from the dataset. Other quality control checks were also performed to exclude images with out of focus regions or large regions of missing tissue. Overall, 422 AF-H&E image pairs and 405 AF-mIF image pairs passed all alignment and quality control checks and were used for model development, while 26 AF-H&E image pairs and 43 AF-mIF image pairs were excluded.

The antigen retrieval requirement of mIF staining is inherently harsher than H&E staining and more likely to create tissue warping, such as minor folds or tears. Therefore, an automatic algorithm to identify local regions with tissue warping was developed to exclude such artifacts. Patches from the globally aligned AF-mIF image pairs at the same location were converted to grayscale and normalized to the [0, 1] range. Using the normalized gradient field (NGF) [25] or normalized total gradient (NTG) [26] as the distance metric between the patches, a translational alignment of the patches was performed using Powell’s method [27] combined with Gaussian pyramids with three levels [28] to speed up the convergence. The magnitude of the translational vector obtained was then used as an alignment score, whereby a lower score indicated better alignment quality. In order to determine a suitable threshold, a small subset of the training slides were manually annotated and compared against the alignment score. As a result, it was determined that any region with an alignment score greater than 6µm at 10× magnification should be considered misaligned and excluded from model development. No significant differences were observed between using NGF or NTG as the distance metric.

#### Data Splits

Cases were randomly assigned to training, validation, and testing splits for virtual stainer model development. The distributions of sex, age, and diagnosis for each individual split were balanced and there were no significant deviations from the overall dataset distribution. For H&E model development, the final dataset consisted of 215 training, 85 validation, and 122 testing slides. For mIF model development, the final dataset consisted of 245 training, 65 validation, and 95 testing slides. A larger number of training slides was allocated for the mIF model development due to the lower prevalence of certain targets, while still maintaining a reasonable number of validation and testing slides.

### Virtual Stainer

#### Overview

Virtual stainer models were developed to generate H&E and mIF virtual stains from unstained AF images. For mIF, four separate models were trained to generate DAPI + PanCK, PD-L1, CD3, and CD8, respectively. All models operated on patches which were then combined into whole slide images (WSIs) for evaluation.

#### Data Sampling

For model development, paired patches of the AF and corresponding stains were sampled from tissue regions of the training and validation slides. Paired patches were sampled at 40× magnification from the same locations and a padding of 16 pixels was added to each side of the corresponding stains to account for local errors in the global alignment algorithm, which was addressed by a shift-invariant regression loss during training and further described in **Loss**. Therefore, the dimensions of the patches were 128×128×20 for AF, 160×160×3 for H&E, and 160×160×1 for each mIF channel.

For the H&E model, patches were uniformly sampled across all tissue regions. Overall, 30 million patches were sampled for training and 1000 patches were sampled for validation. The smaller number of patches chosen for validation was observed to produce comparable metrics with decreased computational times.

For the mIF models, local regions with tissue warping, as determined by the automatic algorithm using the NGF distance metric described in **Data Alignment and Quality Control**, were excluded from sampling. Patches from tumor regions, as determined by the PanCK stain, were sampled with higher probabilities. Overall, for the DAPI + PanCK, CD3, and CD8 models, 10 million patches were sampled for training and 1000 patches were sampled for validation. For PD-L1, a larger number of patches were sampled for training and patches from PD-L1 positive regions, as determined by the intensity of the PD-L1 stain, were also sampled with higher probabilities. Due to the heterogeneity of PD-L1 expression in both immune cells and tumor cells, this was found to improve overall model performance. Overall, for the PD-L1 model, 20 million patches were sampled for training and 1000 patches were sampled for validation.

#### Pseudo-IHC

Pseudo-IHC (pIHC) refers to IHC-like images rendered from mIF images using an algorithm based on modeling absorption using the Beer-Lambert law [29,30]. Our implementation of the algorithm included two modifications to improve the visual quality of the pIHC images. First, a realistic tissue background was rendered using the unmixed residual AF signal, which adds tissue morphology details to the image. Second, color offsets were added to match the colors typically observed in real IHC images obtained using brightfield microscopy, such as in regions where no tissue is present.

Three channels from the mIF image were used to render the corresponding pIHC image. First, the DAPI stain was used to render the corresponding hematoxylin stain, which labels nuclei blue-purple. Second, the target stain was used to render the corresponding 3,3’-diaminobenzidine (DAB) stain, which labels the target brown. Third, the unmixed residual AF was used to render the corresponding tissue background. **Figure 1A** shows examples of the mIF image and the corresponding pIHC image for different targets.

**Figure 1:**
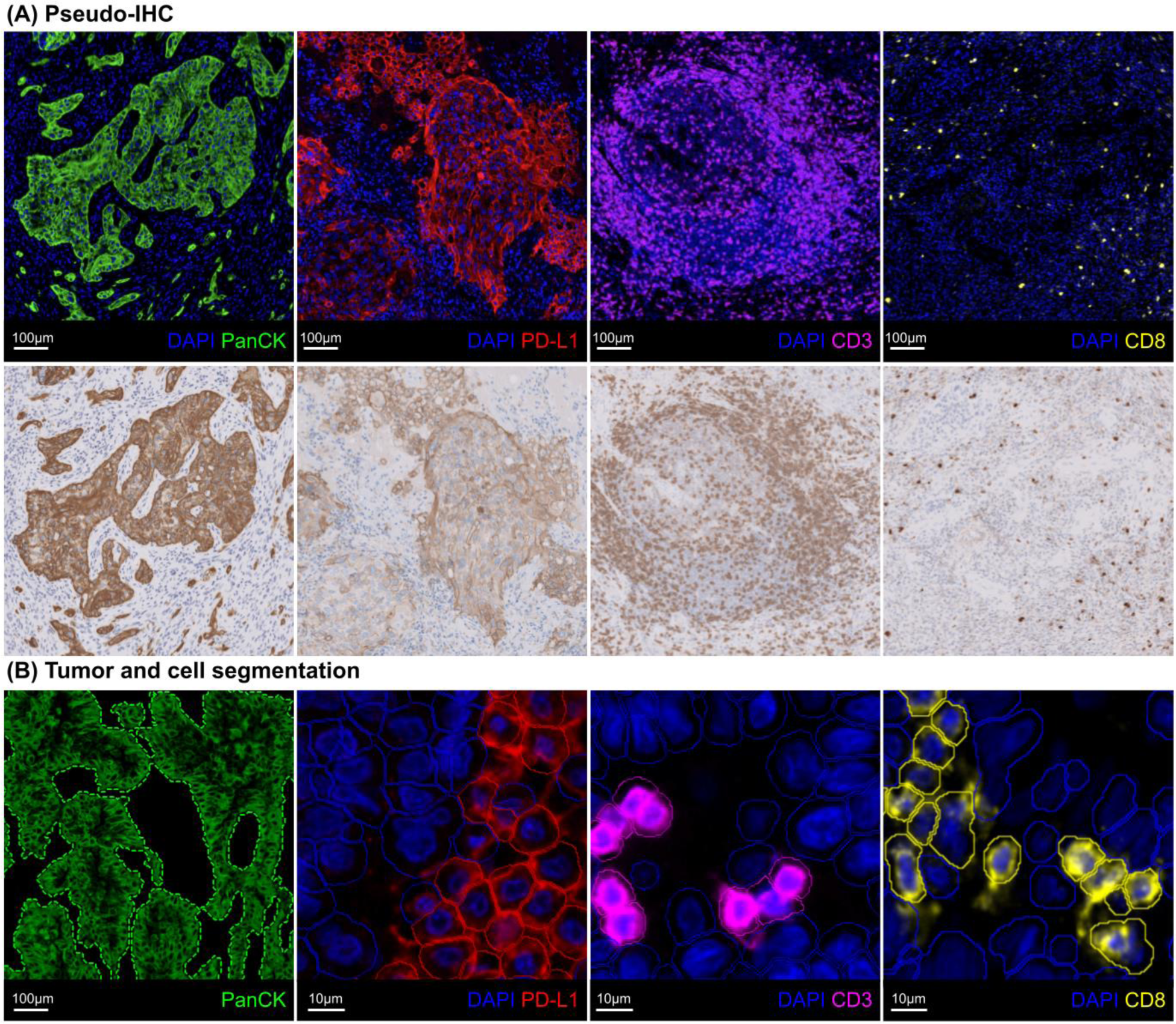
Pseudo-IHC (pIHC) and segmentation algorithms were developed for mIF model development and evaluation. **(A)** Examples of mIF (top) and the corresponding pIHC (bottom) for PanCK, PD-L1, CD3, and CD8. As per conventional practice, the residual unmixed AF is not shown in the mIF to maintain appropriate visual quality. **(B)** Example of the segmentations of tumor regions based on the PanCK stain (dashed outline, green), and negative (solid outline, blue) and positive (solid outline, respective color) cells for PD-L1, CD3, and CD8.

Note that the pIHC algorithm described here is based on pixel-wise image processing, invertible, and can also be used to render the mIF image from the corresponding pIHC image. More details are available in the **Supplementary Material 1.**

Initial experiments were conducted to compare the effectiveness of training the mIF models to predict the output as mIF or pIHC virtual stains. The initial experiments showed that training the models to predict pIHC virtual stains, and using the inverse pIHC algorithm to render the corresponding mIF virtual stains, resulted in higher quality virtual stains, compared to training the models to directly predict mIF virtual stains. This was determined by qualitative review of validation patches and WSIs. Using pIHC also enabled a more direct translation of some of the H&E model hyperparameters to the mIF models.

#### Architecture

The architecture of the models was based on generative adversarial networks (GANs), specifically the ‘pix2pix’ paired image-to-image translation approach, and has been previously described [19,22,31]. Briefly, the architecture consisted of a U-Net-based generator, a conditional discriminator, and two unconditional discriminators which operated at different magnifications of the image. The generator received the AF as the input and was trained to predict the corresponding virtual stain as the output. The discriminators received the real and virtual stains as the input and were trained to differentiate between the real and virtual stains. The unconditional discriminators received only the real and virtual stains as the input, whereas the conditional discriminator also received the AF as the input.

Specific hyperparameters such as kernel sizes, numbers of kernels, dropout rate, and attention gates [32] were different and empirically determined for the H&E and mIF models. More details are available in the **Supplementary Material 2**.

#### Loss

The loss components used to train the models have been previously described [19,22]. Briefly, a shift-invariant regression loss was used to minimize the L1 and L2 errors between the real and virtual stains. Conditional and unconditional adversarial losses were used in a minimax game to force the generator to produce realistic images in order to fool the discriminators. A rotational consistency loss was used to make the output rotation invariant and prevent the model from learning any orientation biases. An L2 regularization loss was used to penalize large model weights and reduce overfitting.

For the mIF models, an additional pixel-wise weighting scheme was applied in the shift-invariant regression loss in order to improve the quality of the target virtual stain, especially for more heterogenous or less prevalent targets, such as PD-L1 and CD8. Specifically, **Equation 1** was used to weight the pixels based on the IF intensity of the target stain when calculating the loss, such that pixels with higher intensities were given more importance. Note that the mIF models were trained to predict pIHC virtual stains as described in **Pseudo-IHC**. Therefore, all losses were calculated using pIHC and the IF intensity of the target stain was used only to derive the weights for the pixel-wise weighting scheme.

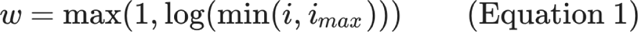

where

*w* = The weight

*i* = The IF intensity of the target stain

*i_max_* = The maximum IF intensity to clip values

For the DAPI + PanCK model, the pixel-wise weighting scheme was not applied, as no significant improvements in quality were observed for the PanCK target stain which is highly prevalent, and the DAPI stain which labels nuclei was also an output of interest. For the CD8 model, all pixels with an IF intensity of the target stain of less than 15 were given a weight of 1, to reduce the effect of label noise from background and non-specific fluorescence which is further described in **Results**.

Specific hyperparameters such as the weights for each loss component were different and empirically determined for the H&E and mIF models. More details are available in **Supplementary Material 2**.

#### Training and Validation

All model weights were randomly initialized using Glorot uniform initialization [33] and optimized using Adam optimization [34] to minimize the total loss on the training set. All models were trained with a fixed batch size. Learning rate schedules were employed to dynamically change the learning rate over time. Loss schedules were employed to change the relative weights of the shift-invariant regression loss and adversarial loss components over time. Performance was monitored based on the L1 error and Frechet Inception Distance (FID) [35] between the real and virtual stain patches to ensure model convergence. The best checkpoint for each model was selected based on a combination of the minimum L1 and FID on validation patches, as well as qualitative review of the virtual stains on validation WSIs. The best checkpoint for all models was between 320,000 and 360,000 training steps.

Specific hyperparameters such as the batch size, learning rate, learning rate schedules, and loss schedules were different and empirically determined for the H&E and mIF models. More details are available in **Supplementary Material 2**.

### Evaluation

Domain-specific computational methods were used to evaluate the accuracy of the H&E and mIF virtual stains compared to the real stains on testing slides.

#### H&E

To evaluate the accuracy of the H&E virtual stains, an independent model which has been previously described [36] was used for automatic histologic subtyping, and the similarity of the model’s segmentations on real and virtual stains was evaluated. Briefly, the model was trained on H&E images of LUAD cases from The Cancer Genome Atlas (TCGA) dataset to segment nine histologic features at 10× magnification. The histologic features include six tumor subtypes (acinar, cribriform, lepidic, micropapillary, papillary, solid), leukocyte aggregates, necrosis, and an “other” category comprising features such as normal tissue and stroma.

The tumor subtypes are only applicable to LUAD cases. Therefore, the six tumor subtypes were combined into a single “combined tumor” category for any analysis involving non-LUAD cases (23 LUSC, 3 adenosquamous carcinoma, 1 undetermined). This was considered a reasonable approach even though the model was not specifically trained for non-LUAD cases, given that it was not the accuracy of the histologic subtyping model that was being evaluated, but rather the similarity of the model’s segmentations on real and virtual stains.

Additionally, each tumor subtype is typically considered clinically-relevant only if its presence exceeds a certain threshold [37]. Therefore, an additional analysis for each tumor subtype was performed by limiting to LUAD cases in which that subtype exceeded 5% of the total tumor area (LUAD-5%). This excluded cases with low prevalence and may better represent clinical performance.

Dice scores for the overlap of segmentations on the real and virtual stains were calculated, whereby higher values indicate better performance.

#### mIF

To evaluate the accuracy of the mIF virtual stains, several cell segmentation-based measurements were obtained using automatic image analysis tools developed using Visiopharm software (Visiopharm A/S, Hoersholm, Denmark). Briefly, cells were identified and segmented based on the DAPI stain and tumor regions were identified and segmented based on the PanCK stain. Positive cell expression of PD-L1, CD3, and CD8 was determined based on the average intensity of the respective stain within each segmented cell. **Figure 1B** shows an example of the segmentations.

Measurements of the positive area, positive cell density, positive cell percentage, and computationally-derived TPS and CPS, were calculated for each stain, where relevant. Specifically, positive area was calculated only for PanCK, while positive cell density and positive cell percentage were calculated only for DAPI, PD-L1, CD3, and CD8. For DAPI, positive cell percentage was not calculated as the calculation is itself a function of the number of cells segmented based on DAPI, and would therefore always be 100%. For PD-L1 only, positive cell percentage was calculated as TPS and CPS to align with clinical terminology and practice. More details are available in the **Supplementary Material 3**.

Additionally, a colocalization analysis which can better represent particular cell subsets was also performed. Specifically, CD3 and CD8 colocalization can better represent the subset of cytotoxic T cells, and CD3 and PD-L1 colocalization can better represent the subset of PD-L1 positive T cells.

Analysis was performed according to three different definitions of the region of interest:

1. **Tissue:** The entire tissue region, as determined by a tissue detection algorithm. The same region was used on both the real and virtual stains. This analysis was useful to measure the performance on the entire WSI.
2. **Real tumor:** The real tumor region, as determined by the real PanCK stain. The same region was used on both the real and virtual stains. This analysis was useful to measure the performance in the tumor region independent of the quality of the virtual PanCK stain.
3. **Respective tumor:** The respective tumor regions, as determined by the real PanCK stain on the real stains and the virtual PanCK stain on the virtual stains. This analysis was useful to measure the performance in the tumor region in a fully-virtual workflow.

Pearson’s correlations and absolute differences between the measurements on real and virtual stains were calculated, whereby higher values for correlations indicate better performance and lower values for absolute differences indicate better performance.

## Results

### H&E

#### Qualitative Analysis

**Figure 2A** shows a WSI example of the H&E real and virtual stains. **Figure 2B** shows a magnification series of concentric regions from the WSI.

**Figure 2:**
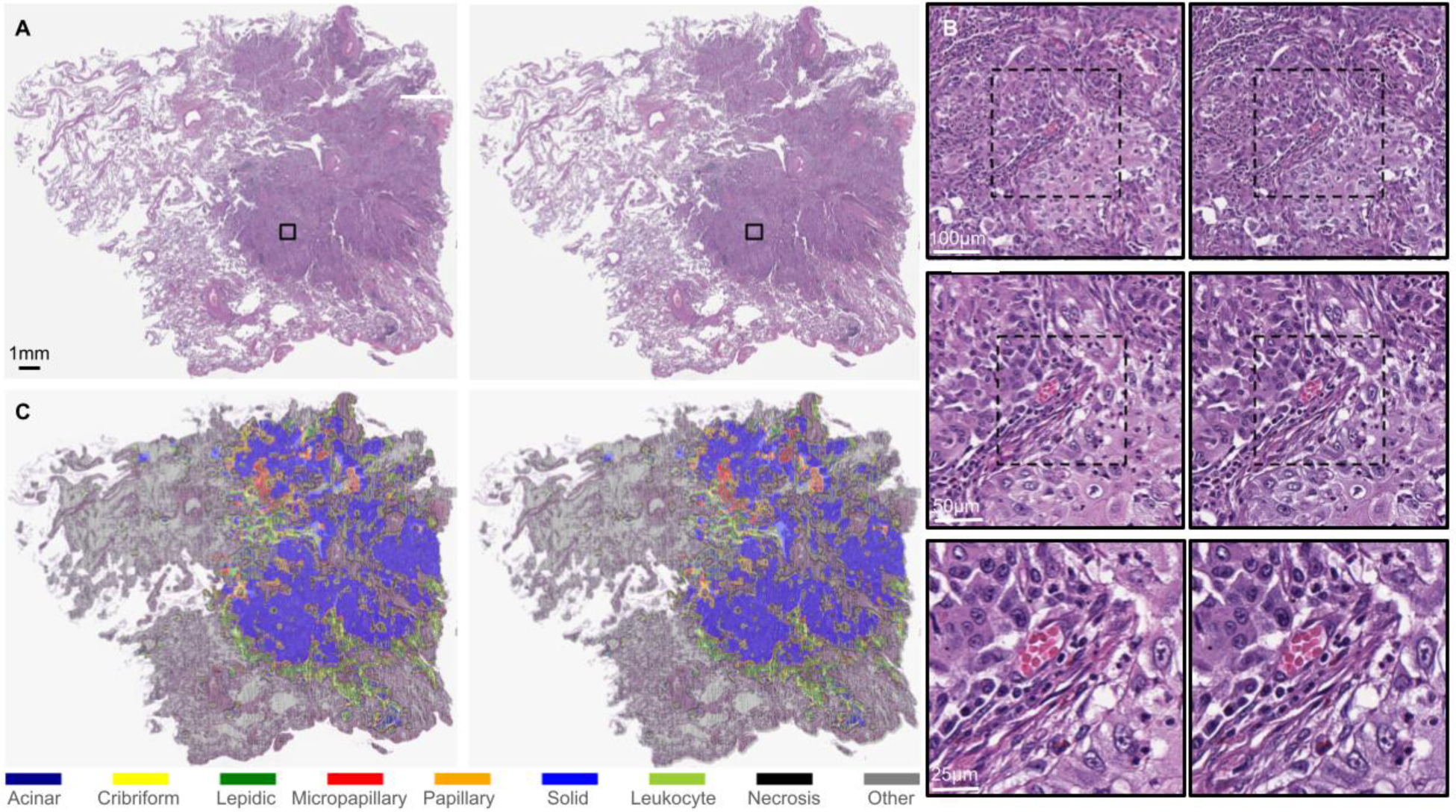
**(A)** Example of real (left) and virtual (right) stains for a H&E WSI. **(B)** Magnification series of real (left) and virtual (right) stains for concentric regions from the WSI at 10× (top), 20× (middle), and 40× (bottom) magnifications. **(C)** Example of the segmentations on real (left) and virtual (right) stains obtained from automatic histologic subtyping.

Overall, the virtual stains were able to reproduce key morphological features at tissue and cell levels such as tumor cells, tumor-infiltrating lymphocytes, and tumor-associated stroma. Certain subcellular features such as cell borders, nuclear size and shape, and cytoplasm appearance, as well as mitoses, nuclear lobation, and nucleoli, were also reproduced in the virtual stains.

#### Quantitative Analysis

**Tables 1** and **2** show the average performance based on the overlap of segmentations obtained from automatic histologic subtyping of real and virtual stains. **Figure 2C** shows examples of the segmentations.

**Table 1:** Average Dice score (mean ± SD, median) between segmentations on real and virtual stains of combined tumor, leukocyte aggregates, necrosis and “other” categories on testing slides.

| Category | All (122 cases) | LUAD (95 cases) | Non-LUAD (27 cases) |
| --- | --- | --- | --- |
| Combined tumor | $0.92 \pm 0.05$,<br>0.94 | $0.93 \pm 0.05$,<br>0.94 | $0.92 \pm 0.04$,<br>0.92 |
| Leukocyte aggregates | $0.85 \pm 0.06$,<br>0.86 | $0.85 \pm 0.06$,<br>0.86 | $0.85 \pm 0.06$,<br>0.85 |
| Necrosis | $0.66 \pm 0.27$,<br>0.76 | $0.62 \pm 0.29$,<br>0.71 | $0.78 \pm 0.13$,<br>0.82 |
| Other | $0.95 \pm 0.04$,<br>0.96 | $0.95 \pm 0.04$,<br>0.97 | $0.94 \pm 0.04$,<br>0.95 |

**Table 2:** Average Dice score (mean ± SD, median) between segmentations on real and virtual stains of six tumor subtypes on LUAD testing slides.

| Tumor Subtype | LUAD (95 cases) | LUAD-5% (Number of cases meeting criteria) |
| --- | --- | --- |
| Acinar | $0.73 \pm 0.17$,<br>0.77 | $0.80 \pm 0.09$,<br>0.82 (59) |
| Cribriform | $0.56 \pm 0.22$,<br>0.63 | $0.70 \pm 0.12$,<br>0.71 (36) |
| Lepidic | $0.76 \pm 0.15$,<br>0.79 | $0.80 \pm 0.09$,<br>0.82 (76) |
| Micropapillary | $0.62 \pm 0.27$,<br>0.69 | $0.82 \pm 0.06$,<br>0.84 (23) |
| Papillary | $0.68 \pm 0.25$,<br>0.77 | $0.81 \pm 0.07$,<br>0.82 (56) |
| Solid | $0.76 \pm 0.14$,<br>0.76 | $0.79 \pm 0.12$,<br>0.80 (76) |

Overall, the segmentations on real and virtual stains were very similar, despite patches often exhibiting multiple subtypes or ambiguous categorization. The segmentations for the combined tumor, leukocyte aggregates, and “other” categories showed the best performance, with Dice scores above 0.8, indicating good differentiation of tumor, non-tumor, and immune cells. The performance for the necrosis category was slightly lower, which may be attributed to low AF signals in necrotic regions. No significant differences were observed between LUAD and non-LUAD cases. For LUAD cases, the Dice scores ranged from moderate (0.5 - 0.7) to good (0.7 - 0.9) for the segmentations of the tumor subtypes. The best performance was observed for the segmentations of solid, acinar, and lepidic tumor subtypes, which are the most established and common tumor subtypes [37]. The lowest performance was observed for the segmentations of the cribriform tumor subtype, which is less established and more difficult to characterize than the other tumor subtypes [38]. The performance for all tumor subtypes improved for the subset of LUAD-5% cases, with all Dice scores above 0.7, demonstrating promising performance when factoring in a clinical threshold.

### mIF

#### Qualitative Analysis

**Figures 3 and 4** show examples of the mIF real and virtual stains. **Figure 3** shows a WSI composite of all mIF stains and the individual model outputs at several regions across the WSI. **Figure 4** shows examples of key immune phenotypes which are important for immuno-oncology. More examples are available in **Supplementary Material 4**.

**Figure 3:**
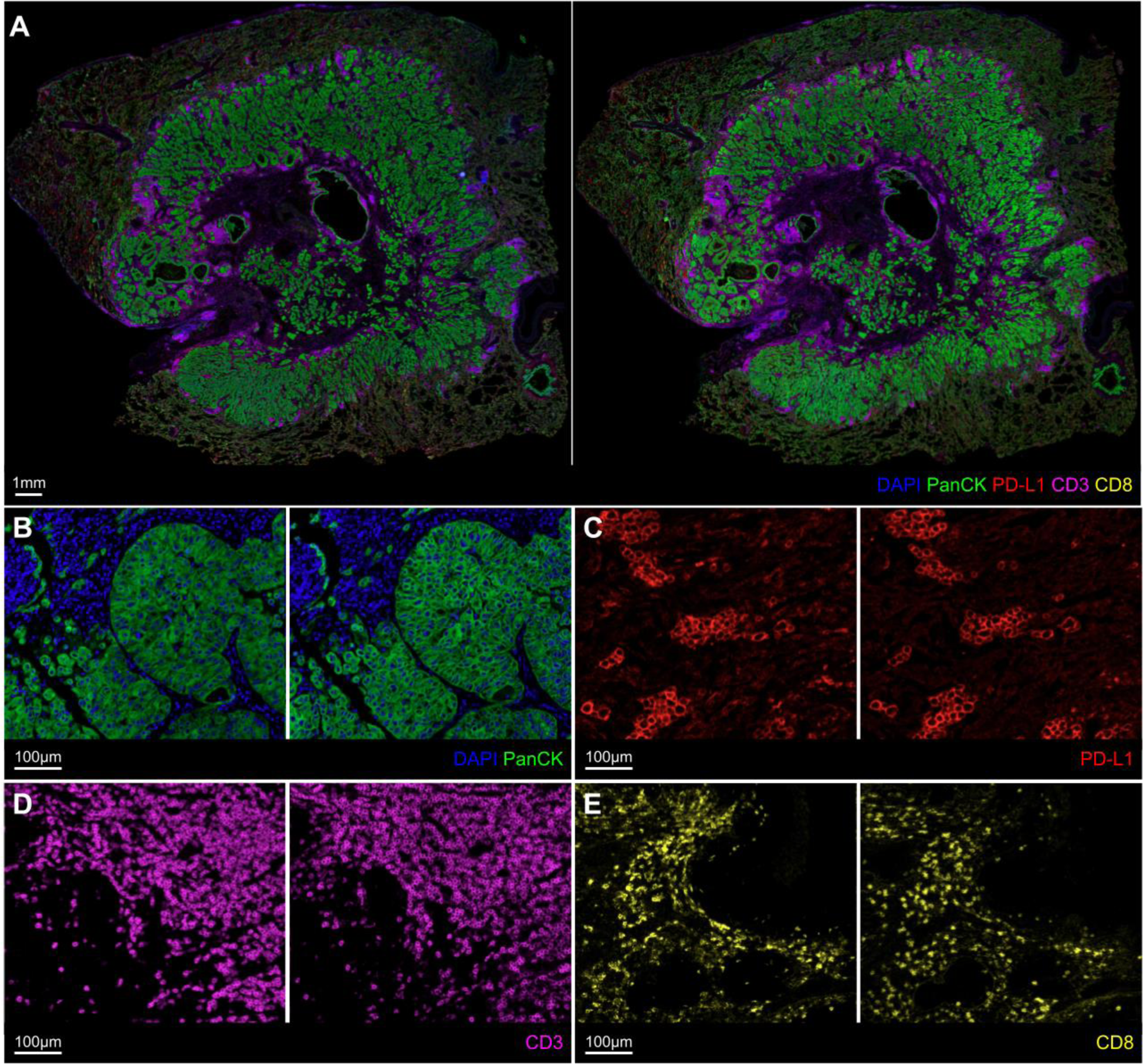
**(A)** Example of real (left) and virtual (right) stains showing the WSI composite for all mIF stains. **(B-E)** Examples of real (left) and virtual (right) stains from the individual models for DAPI + PanCK, PD-L1, CD3, and CD8 at several regions across the WSI.

**Figure 4:**
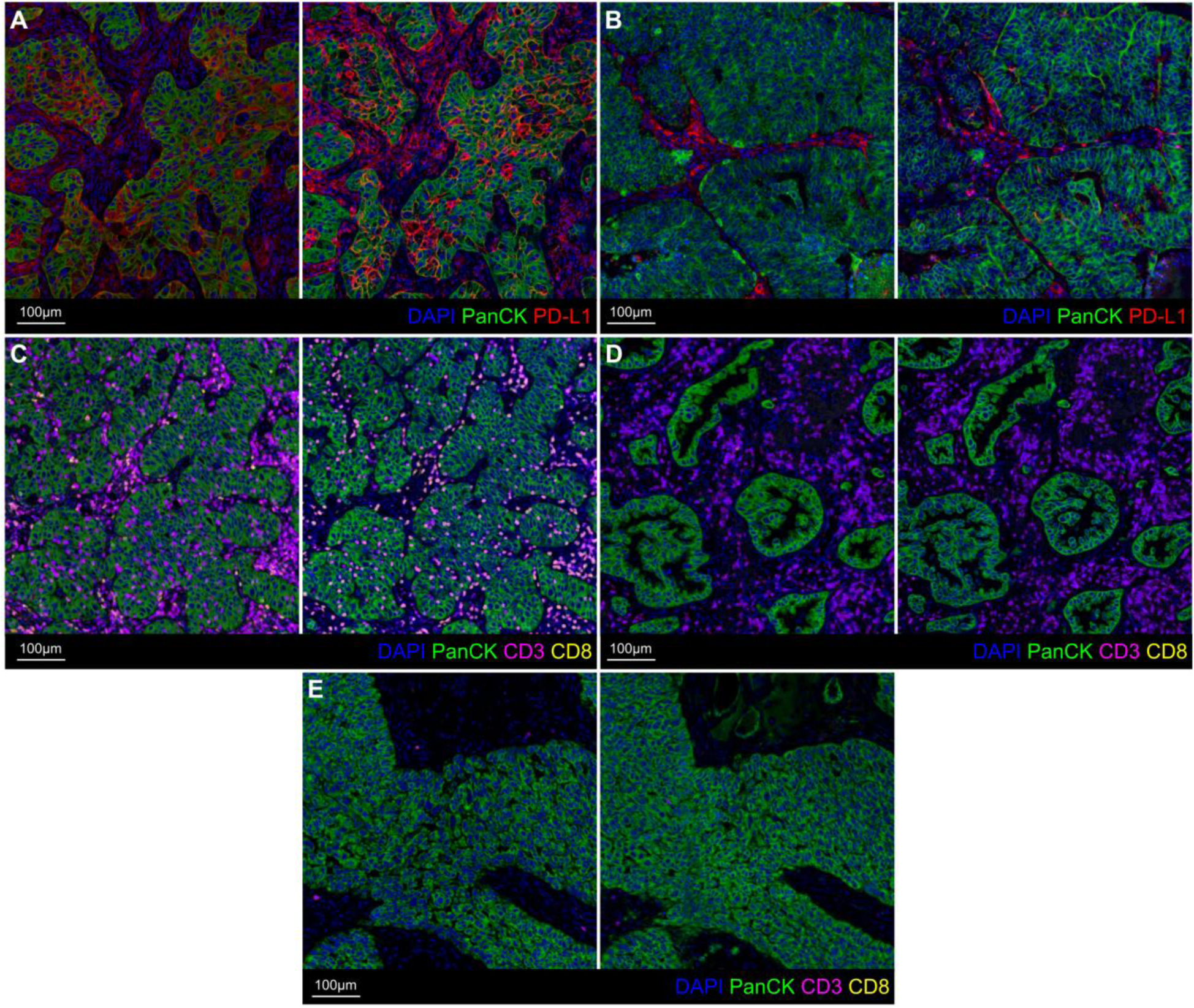
Examples of real (left) and virtual (right) stains showing key immune phenotypes. **(A)** PD-L1 positive tumor. **(B)** PD-L1 negative tumor. **(C)** Immune inflamed tumor with T cell infiltration. **(D)** Immune excluded tumor with T cell accumulation in surrounding stroma. **(E)** Immune desert tumor with no T cells.

Overall, the global distribution of the virtual stains were similar to the real stains. The virtual stains were able to satisfactorily reproduce key morphological features and protein biomarker expressions at tissue and cell levels. Importantly, the identification of key immune phenotypes such as PD-L1 positive, PD-L1 negative, immune inflamed, immune excluded, and immune desert tumors was attainable with the virtual stains [12]. The virtual stains for morphological or cell type biomarkers such as DAPI, PanCK, and CD3 were of higher quality and accuracy than highly dynamic cell state biomarkers such as PD-L1. For CD8, while the overall spatial distribution and relative density of positive cells were moderately accurate at the tissue level, the positive expression at cell level was not as accurate. Upon further inspection, high amounts of background and non-specific fluorescence were observed in many of the CD8 real stains, which adds label noise during the model training procedure, resulting in poorer performance. More details are available in **Supplementary Material 4**.

#### Quantitative Analysis

**Tables 3** and **4** show the Pearson’s correlations between the measurements on real and virtual stains obtained from the cell segmentation-based analysis in Visiopharm software for the single expression and colocalization analysis, respectively. Blank entries indicate measurements that were not relevant for the stain as described in **Materials and Methods**. The average absolute differences and scatterplots of the measurements on real and virtual stains are available in **Supplementary Material 4**.

**Table 3:** Pearson’s correlation (95% CI) between measurements on real and virtual stains obtained from the cell segmentation-based analysis in Visiopharm software for PanCK, DAPI, PD-L1, CD3, and CD8 on testing slides. Analysis was performed according to three different definitions of the region of interest.

| Region | Measurement | PanCK | DAPI | PD-L1 | CD3 | CD8 |
| --- | --- | --- | --- | --- | --- | --- |
| Tissue | Positive area (mm<sup>2</sup>) | 0.96<br>(0.94, 0.97) | - | - | - | - |
|  | Positive cell density (cells/mm<sup>2</sup>) | - | 0.85<br>(0.79, 0.90) | 0.63<br>(0.49, 0.74) | 0.89<br>(0.83, 0.92) | 0.41<br>(0.23, 0.56) |
|  | Positive cell percentage (%) | - | - | 0.50<br>(0.33, 0.64) | 0.84<br>(0.77, 0.89) | 0.43<br>(0.25, 0.58) |
| Real tumor | Positive cell density (count/mm<sup>2</sup>) | - | 0.66<br>(0.52, 0.76) | 0.52<br>(0.36, 0.66) | 0.84<br>(0.76, 0.89) | 0.50<br>(0.33, 0.64) |
|  | Positive cell percentage (%) | - | - | See<br>TPS and CPS | 0.79<br>(0.70, 0.86) | 0.45<br>(0.27, 0.60) |
|  | TPS (%) | - | - | 0.46<br>(0.29, 0.61) | - | - |
|  | CPS (%) | - | - | 0.71<br>(0.60, 0.80) | - | - |
| Respective tumor | Positive cell density (cells/mm<sup>2</sup>) | - | 0.53<br>(0.37, 0.66) | 0.52<br>(0.36, 0.66) | 0.84<br>(0.76, 0.89) | 0.49<br>(0.32, 0.63) |
|  | Positive cell percentage (%) | - | - | See<br>TPS and CPS | 0.76<br>(0.66, 0.83) | 0.45<br>(0.28, 0.60) |
|  | TPS (%) | - | - | 0.47<br>(0.29, 0.61) | - | - |
|  | CPS (%) | - | - | 0.71<br>(0.59, 0.80) | - | - |

**Table 4:** Pearson’s correlation (95% CI) between measurements on real and virtual stains obtained from the colocalization analysis in Visiopharm software for CD3 and CD8, and CD3 and PD-L1, on testing slides. Analysis was performed according to three different definitions of the region of interest.

| Region | Measurement | CD3 and CD8 | CD3 and PD-L1 |
| --- | --- | --- | --- |
| Tissue | Positive cell density (cells/mm<sup>2</sup>) | 0.53<br>(0.38, 0.66) | 0.81<br>(0.73, 0.87) |
|  | Positive cell percentage (%) | 0.55<br>(0.39, 0.67) | 0.73<br>(0.62, 0.81) |
| Real tumor | Positive cell density (cells/mm<sup>2</sup>) | 0.56<br>(0.41, 0.69) | 0.71<br>(0.60, 0.80) |
|  | Positive cell percentage (%) | 0.56<br>(0.41, 0.69) | 0.66<br>(0.53, 0.76) |
| Respective tumor | Positive cell density (cells/mm<sup>2</sup>) | 0.51<br>(0.35, 0.65) | 0.75<br>(0.65, 0.83) |
|  | Positive cell percentage (%) | 0.53<br>(0.36, 0.66) | 0.68<br>(0.55, 0.77) |

Overall, the correlations ranged from moderate (0.4 - 0.7) to good (0.7 - 1.0) for the different measurements. The performance metrics were consistent with the qualitative observations described above, whereby virtual stains for DAPI, PanCK, and CD3 showed better performance than PD-L1 and CD8. In general, there was a decrease in performance in tumor regions compared to tissue regions, whereby the correlations were lower in tumor regions, indicating that biomarker expressions in tumor regions may be more dysregulated or heterogeneous and difficult to predict. For PD-L1, the correlations between the measurements on real and virtual stains were 0.46 for TPS and 0.71 for CPS, compared to previously reported inter-observer agreements (Cohen’s kappa) of 0.53 - 0.72 for TPS [39–41] and 0.52 - 0.74 for CPS [39] across various studies. The measurements of CPS showed better correlations than TPS, indicating that PD-L1 expression can be predicted more accurately in immune cells than in tumor cells. This was consistent with the good performance of the CD3 and PD-L1 colocalization analysis, which represents PD-L1 expression on T cells. For CD8, the background autofluorescence and non-specific fluorescence contributed to the false positive identification of cells. While CD8 can be expressed on natural killer cells and dendritic cells as well, the primary cell type of interest when staining for CD8 is often cytotoxic T cells. Therefore, the colocalization analysis of CD3 and CD8 enabled the specific analysis of cytotoxic T cells. The high quality of both the real and virtual stains for CD3 also reduced the false positives and improved the performance metrics.

## Discussion

In this study, we present virtual stainer models that can generate virtual stains for H&E and an immuno-oncology mIF panel (DAPI, PanCK, PD-L1, CD3, CD8) from AF images of unstained NSCLC tissue specimens. This work follows our previous work [19,22] and demonstrates the generalizability of our technique by extending its application to a different disease and stain modality.

The H&E virtual stains accurately reproduced key morphological features at tissue, cell, and even subcellular levels, compared to real stains. The virtual stains also demonstrated good performance when used for automatic histologic subtyping. Overall, there was good differentiation between tumor and non-tumor regions, with Dice scores above 0.8. There was also good characterization of immune cells, with Dice scores of 0.85 for leukocyte aggregates. For LUAD cases, the differentiation between specific tumor subtypes also showed good performance, especially for the solid, acinar, and lepidic tumor subtypes, with Dice scores above 0.7. Therefore, the H&E virtual stains showed potential to be accurate enough for the diagnosis and subtyping of NSCLC cases, as it is routinely used for in clinical practice. We hypothesize that some of the differences between the segmentations on real and virtual stains may be partly attributed to the variability in pathologist annotations used for training the histologic subtyping model [36], whereby the high rates of inter-pathologist disagreement observed when subtyping adenocarcinoma [42] may result in the histologic subtyping model being susceptible to minor and clinically-irrelevant variations between the real and virtual stains.

The mIF virtual stains also showed good performance, reproducing key morphological features and protein biomarker expressions at tissue and cell levels, especially for DAPI, PanCK, and CD3. The identification of key immune phenotypes important for immuno-oncology was also attainable with the virtual stains. Overall, there was good agreement between clinically-relevant measurements derived from the real and virtual stains, especially for measurements of the tumor area, T cell density and percentage, and CPS, with correlations above 0.7. For PD-L1, the performance of the virtual stains was better in immune cells than in tumor cells, and further investigation would be required to determine the reason for this difference. We hypothesize that the more distinct morphology of immune cells, the heterogeneity of dysregulated PD-L1 expression in tumor cells, as well as the difference in AF signals between immune cells and tumor cells, partly contribute to this difference. For CD8, we observed background and non-specific fluorescence in the real stains which adds label noise during the model training procedure. Therefore, we expect the performance of the CD8 virtual stains to considerably improve with a less noisy dataset, which can be obtained by using fluorophores at a different wavelength for CD8 staining, for example, and better quality control. We aim to further improve on the performance of PD-L1 and CD8 virtual stains in future work.

We also present a pIHC algorithm which can be used to render IHC-like images from mIF images, introducing modifications to existing methods [29,30] to improve the visual quality and similarity of pIHC images to real IHC images that pathologists are more familiar with reading and evaluating. The pIHC algorithm can be used in several ways. First, it enables the generation of pIHC images for multiple biomarkers from a single mIF image. On the other hand, the generation of real IHC images for multiple biomarkers would require the preparation of multiple slides containing consecutive sections obtained from a tissue block, which can be limited by the availability of tissue. The use of consecutive sections also prohibits accurate colocalization analysis of biomarkers, which is required for specific cell subsets and spatial analysis. Second, it enables flexibility in the choice of modality for different applications. For example, pIHC images may be preferred when pathologists review images, whereas mIF images may be preferred for the development of computational analysis workflows and spatial biology research. We also took advantage of this flexibility in our model development process, whereby using pIHC enabled a more direct translation of previous virtual stainer models that were developed for H&E and IHC images, thereby reducing model development and iteration time. Similarly, many existing pathology image analysis models may be developed for IHC images and the use of pIHC may directly enable mIF images to be applied to those models.

In this study, the accuracy of the virtual stains compared to the real stains was evaluated primarily using computational methods. While results were promising, further validation would be required to establish if the virtual stains can be used in clinical or research settings as a substitute for real stains. Human reader studies should be performed to evaluate the diagnostic accuracy of virtual stains by pathologists. Previous reader studies have shown that H&E virtual stains can be accurate enough for pathologists to grade images using established clinical scoring systems in nonalcoholic steatohepatitis [19] and prostate cancer [22]. A similar reader study can be performed for H&E virtual stains in NSCLC based on histologic classification and subtyping. While mIF image analysis is often computational and there are few established scoring systems based on mIF for NSCLC, more in-depth computational spatial analysis comparing the real and virtual stains can be performed for further validation. For example, more advanced machine learning-based algorithms [43] could be developed to improve upon the current threshold-based algorithms used for cell segmentation, which can have limited accuracy in separating overlapping adjacent cells. Additionally, it would be important to evaluate the association with clinical outcomes or endpoints, in order to determine any clinical impact of the differences observed between the measurements on real and virtual stains.

In summary, we demonstrate the feasibility of generating virtual stains for H&E and mIF from AF images of unstained NSCLC tissue specimens, extending the application of our previous work to another disease and stain modality. Virtual staining has great potential to enable spatial biology research, improve efficiency and reliability in the clinical workflow, as well as conserve tissue samples in a non-destructive manner for future analysis. This study reinforces the potential of digital pathology and virtual staining to improve medical decision making and patient outcomes.

## Data Availability

Due to the nature of this research, data is not available.

## Acknowledgments

We thank Verily and Genmab for supporting this collaboration. We thank Roopam Rajvanshi, Sebastian Dobon, Bryan Crampton, and Pavithran Ramachandran for data infrastructure support. We thank Byron Bogaert and Amer Jarrah for computer hardware and network support. We thank Janelle Chang Clark, Susan Kram, and Tyler Hassenpflug for program management and operations support. We thank Melissa Miao and Radha Patel for business development support. We thank Fabien Beckers for program leadership support.

## Abbreviations

**AF**, autofluorescence

**CD3**, cluster of differentiation 3

**CD8**, cluster of differentiation 8

**CI**, confidence interval

**CPS**, combined positive score

**DAB**, 3,3’-diaminobenzidine

**DAPI**, 4′,6-diamidino-2-phenylindole

**FFPE**, formalin-fixed paraffin-embedded

**FID**, Frechet Inception Distance

**GAN**, generative adversarial network

**H&E**, hematoxylin and eosin

**HER2**, human epidermal growth factor receptor 2

**IHC**, immunohistochemistry

**LUAD**, lung adenocarcinoma

**LUSC**, lung squamous cell carcinoma

**mIF**, multiplex immunofluorescence

**NGF**, normalized gradient field

**NSCLC**, non-small cell lung cancer

**NTG**, normalized total gradient

**PanCK**, pancytokeratin

**PD-1**, programmed cell death 1

**PD-L1**, programmed cell death ligand 1

**pIHC**, pseudo-immunohistochemistry

**SD**, standard deviation

**TCGA**, The Cancer Genome Atlas

**TPS**, tumor proportion score

**WSI**, whole slide image.

## References

1. Gridelli C, Rossi A, Carbone DP, Guarize J, Karachaliou N, Mok T, et al. Non-small-cell lung cancer. Nat Rev Dis Primers. 2015;1: 15009.

2. Herbst RS, Morgensztern D, Boshoff C. The biology and management of non-small cell lung cancer. Nature. 2018;553: 446–454.

3. Harms PW, Frankel TL, Moutafi M, Rao A, Rimm DL, Taube JM, et al. Multiplex Immunohistochemistry and Immunofluorescence: A Practical Update for Pathologists. Mod Pathol. 2023;36: 100197.

4. Francisco-Cruz A, Parra ER, Tetzlaff MT, Wistuba II. Multiplex Immunofluorescence Assays. In: Thurin M, Cesano A, Marincola FM, editors. Biomarkers for Immunotherapy of Cancer: Methods and Protocols. New York, NY: Springer New York; 2020. pp. 467–495.

5. Rojas F, Hernandez S, Lazcano R, Laberiano-Fernandez C, Parra ER. Multiplex Immunofluorescence and the Digital Image Analysis Workflow for Evaluation of the Tumor Immune Environment in Translational Research. Front Oncol. 2022;12: 889886.

6. Teixidó C, Vilariño N, Reyes R, Reguart N. PD-L1 expression testing in non-small cell lung cancer. Ther Adv Med Oncol. 2018;10: 1758835918763493.

7. De Marchi P, Leal LF, Duval da Silva V, da Silva ECA, Cordeiro de Lima VC, Reis RM. PD-L1 expression by Tumor Proportion Score (TPS) and Combined Positive Score (CPS) are similar in non-small cell lung cancer (NSCLC). J Clin Pathol. 2021;74: 735–740.

8. Herbst RS, Baas P, Kim D-W, Felip E, Pérez-Gracia JL, Han J-Y, et al. Pembrolizumab versus docetaxel for previously treated, PD-L1-positive, advanced non-small-cell lung cancer (KEYNOTE-010): a randomised controlled trial. Lancet. 2016;387: 1540–1550.

9. Reck M, Rodríguez-Abreu D, Robinson AG, Hui R, Csőszi T, Fülöp A, et al. Pembrolizumab versus Chemotherapy for PD-L1-Positive Non-Small-Cell Lung Cancer. N Engl J Med. 2016;375: 1823–1833.

10. Mok TSK, Wu Y-L, Kudaba I, Kowalski DM, Cho BC, Turna HZ, et al. Pembrolizumab versus chemotherapy for previously untreated, PD-L1-expressing, locally advanced or metastatic non-small-cell lung cancer (KEYNOTE-042): a randomised, open-label, controlled, phase 3 trial. Lancet. 2019;393: 1819–1830.

11. Kulangara K, Zhang N, Corigliano E, Guerrero L, Waldroup S, Jaiswal D, et al. Clinical Utility of the Combined Positive Score for Programmed Death Ligand-1 Expression and the Approval of Pembrolizumab for Treatment of Gastric Cancer. Arch Pathol Lab Med. 2019;143: 330–337.

12. Tiwari A, Oravecz T, Dillon LA, Italiano A, Audoly L, Fridman WH, et al. Towards a consensus definition of immune exclusion in cancer. Front Immunol. 2023;14: 1084887.

13. Paijens ST, Vledder A, de Bruyn M, Nijman HW. Tumor-infiltrating lymphocytes in the immunotherapy era. Cell Mol Immunol. 2021;18: 842–859.

14. Hermitte F. Biomarkers immune monitoring technology primer: Immunoscore® Colon. J Immunother Cancer. 2016;4: 57.

15. Angell HK, Bruni D, Barrett JC, Herbst R, Galon J. The Immunoscore: Colon Cancer and Beyond. Clin Cancer Res. 2020;26: 332–339.

16. Anagnostou VK, Welsh AW, Giltnane JM, Siddiqui S, Liceaga C, Gustavson M, et al. Analytic variability in immunohistochemistry biomarker studies. Cancer Epidemiol Biomarkers Prev. 2010;19: 982–991.

17. Xu Z, Reilley M, Li R, Xu M. Mapping absolute tissue endogenous fluorophore concentrations with chemometric wide-field fluorescence microscopy. J Biomed Opt. 2017;22: 66009.

18. Deal J, Harris B, Martin W, Lall M, Lopez C, Rider P, et al. Demystifying autofluorescence with excitation scanning hyperspectral imaging. Imaging, Manipulation, and Analysis of Biomolecules, Cells, and Tissues XVI. SPIE; 2018. pp. 129–136.

19. McNeil C, Wong PF, Sridhar N, Wang Y, Santori C, Wu C-H, et al. An End-to-End Platform for Digital Pathology Using Hyperspectral Autofluorescence Microscopy and Deep Learning-Based Virtual Histology. Mod Pathol. 2024;37: 100377.

20. Bai B, Yang X, Li Y, Zhang Y, Pillar N, Ozcan A. Deep learning-enabled virtual histological staining of biological samples. Light Sci Appl. 2023;12: 57.

21. Bai B, Wang H, Li Y, de Haan K, Colonnese F, Wan Y, et al. Label-Free Virtual HER2 Immunohistochemical Staining of Breast Tissue using Deep Learning. BME Front. 2022;2022: 9786242.

22. Wong PF, McNeil C, Wang Y, Paparian J, Santori C, Gutierrez M, et al. Clinical-Grade Validation of an Autofluorescence Virtual Staining System with Human Experts and a Deep Learning System for Prostate Cancer. medRxiv. 2024. p. 2024.03.27.24304447. doi:10.1101/2024.03.27.24304447

23. Burlingame EA, McDonnell M, Schau GF, Thibault G, Lanciault C, Morgan T, et al. SHIFT: speedy histological-to-immunofluorescent translation of a tumor signature enabled by deep learning. Sci Rep. 2020;10: 17507.

24. Ghahremani P, Li Y, Kaufman A, Vanguri R, Greenwald N, Angelo M, et al. Deep Learning-Inferred Multiplex ImmunoFluorescence for Immunohistochemical Image Quantification. Nat Mach Intell. 2022;4: 401–412.

25. Haber E, Modersitzki J. Intensity Gradient Based Registration and Fusion of Multi-modal Images. Medical Image Computing and Computer-Assisted Intervention – MICCAI 2006. Springer Berlin Heidelberg; 2006. pp. 726–733.

26. Shu-Jie Chen, Hui-Liang Shen, Chunguang Li, Xin JH. Normalized Total Gradient: A New Measure for Multispectral Image Registration. IEEE Trans Image Process. 2018;27: 1297–1310.

27. Powell MJD. An efficient method for finding the minimum of a function of several variables without calculating derivatives. Comput J. 1964;7: 155–162.

28. Adelson EH, Anderson CH, Bergen JR, Burt PJ, Ogden JM. Pyramid Methods in Image Processing. RCA Engineer. 1984;29: 33–41.

29. Giacomelli MG, Husvogt L, Vardeh H, Faulkner-Jones BE, Hornegger J, Connolly JL, et al. Virtual Hematoxylin and Eosin Transillumination Microscopy Using Epi-Fluorescence Imaging. PLoS One. 2016;11: e0159337.

30. Yoshitake T, Giacomelli MG, Quintana LM, Vardeh H, Cahill LC, Faulkner-Jones BE, et al. Rapid histopathological imaging of skin and breast cancer surgical specimens using immersion microscopy with ultraviolet surface excitation. Sci Rep. 2018;8: 4476.

31. Isola P, Zhu J-Y, Zhou T, Efros AA. Image-to-image translation with conditional adversarial networks. 2017 IEEE Conference on Computer Vision and Pattern Recognition (CVPR). IEEE; 2017. doi:10.1109/cvpr.2017.632

32. Oktay O, Schlemper J, Le Folgoc L, Lee M, Heinrich M, Misawa K, et al. Attention U-Net: Learning Where to Look for the Pancreas. arXiv [cs.CV]. 2018. doi:10.7937/K9/TCIA.2016.tNB1kqBU

33. Glorot X, Bengio Y. Understanding the difficulty of training deep feedforward neural networks. AISTATS. 2010; 249–256.

34. Kingma DP, Ba J. Adam: A Method for Stochastic Optimization. arXiv [cs.LG]. 2014. Available: http://arxiv.org/abs/1412.6980

35. Heusel M, Ramsauer H, Unterthiner T, Nessler B, Hochreiter S. GANs trained by a two time-scale update rule converge to a local Nash equilibrium. Adv Neural Inf Process Syst. 2017; 6626–6637.

36. Sadhwani A, Chang H-W, Behrooz A, Brown T, Auvigne-Flament I, Patel H, et al. Comparative analysis of machine learning approaches to classify tumor mutation burden in lung adenocarcinoma using histopathology images. Sci Rep. 2021;11: 16605.

37. Travis WD, Brambilla E, Noguchi M, Nicholson AG, Geisinger KR, Yatabe Y, et al. International association for the study of lung cancer/american thoracic society/european respiratory society international multidisciplinary classification of lung adenocarcinoma. J Thorac Oncol. 2011;6: 244–285.

38. Warth A, Muley T, Kossakowski C, Stenzinger A, Schirmacher P, Dienemann H, et al. Prognostic impact and clinicopathological correlations of the cribriform pattern in pulmonary adenocarcinoma. J Thorac Oncol. 2015;10: 638–644.

39. Noske A, Wagner D-C, Schwamborn K, Foersch S, Steiger K, Kiechle M, et al. Interassay and interobserver comparability study of four programmed death-ligand 1 (PD-L1) immunohistochemistry assays in triple-negative breast cancer. Breast. 2021;60: 238–244.

40. Cooper WA, Russell PA, Cherian M, Duhig EE, Godbolt D, Jessup PJ, et al. Intra- and Interobserver Reproducibility Assessment of PD-L1 Biomarker in Non-Small Cell Lung Cancer. Clin Cancer Res. 2017;23: 4569–4577.

41. van Eekelen L, Spronck J, Looijen-Salamon M, Vos S, Munari E, Girolami I, et al. Comparing deep learning and pathologist quantification of cell-level PD-L1 expression in non-small cell lung cancer whole-slide images. Sci Rep. 2024;14: 7136.

42. Thunnissen E, Beasley MB, Borczuk AC, Brambilla E, Chirieac LR, Dacic S, et al. Reproducibility of histopathological subtypes and invasion in pulmonary adenocarcinoma. An international interobserver study. Mod Pathol. 2012;25: 1574–1583.

43. Caicedo JC, Roth J, Goodman A, Becker T, Karhohs KW, Broisin M, et al. Evaluation of Deep Learning Strategies for Nucleus Segmentation in Fluorescence Images. Cytometry A. 2019;95: 952–965.

44. Loshchilov I, Hutter F. SGDR: Stochastic Gradient Descent with Warm Restarts. arXiv [cs.LG]. 2016. Available: http://arxiv.org/abs/1608.03983

