## Supplementary Material for "Autofluorescence Virtual Staining System for H&E Histology and Multiplex Immunofluorescence Applied to Immuno-Oncology Biomarkers in Lung Cancer"

#### 1. Pseudo-IHC

To render the corresponding pIHC image from the mIF image, **Equation S1** was applied to all pixels in the image. Note that **Equation S1** is invertible and can also be used to render the mIF image from the corresponding pIHC image.

$$\begin{bmatrix} IHC_{\mathrm{r}} \\ IHC_{\mathrm{g}} \\ IHC_{\mathrm{b}} \end{bmatrix} = \begin{bmatrix} B_{\mathrm{r}} \\ B_{\mathrm{g}} \\ B_{\mathrm{b}} \end{bmatrix} + \left( \begin{bmatrix} W_{\mathrm{r}} \\ W_{\mathrm{g}} \\ W_{\mathrm{b}} \end{bmatrix} - \begin{bmatrix} B_{\mathrm{r}} \\ B_{\mathrm{g}} \\ B_{\mathrm{b}} \end{bmatrix} \right) \exp\left( - \begin{bmatrix} \alpha_{\mathrm{dapi,r}} & \alpha_{\mathrm{target,r}} & \alpha_{\mathrm{residual,r}} \\ \alpha_{\mathrm{dapi,g}} & \alpha_{\mathrm{target,g}} & \alpha_{\mathrm{residual,g}} \\ \alpha_{\mathrm{dapi,b}} & \alpha_{\mathrm{target,b}} & \alpha_{\mathrm{residual,b}} \end{bmatrix} \begin{bmatrix} IF_{\mathrm{dapi}} \\ IF_{\mathrm{target}} \\ IF_{\mathrm{residual}} \end{bmatrix} \right) \text{(Equation S1)}$$

where

$$\begin{aligned} \mathrm{r, g, b} &= \text{The red, green, and blue channels} \\ B &= \text{The black offset color} \\ W &= \text{The white offset color} \\ \alpha &= \text{The absorption coefficient} \\ IHC &= \text{The IHC pixel value} \\ IF &= \text{The IF pixel value} \end{aligned}$$

To avoid oversaturation of the tissue background, $IF_{\mathrm{residual}}$ was clipped to a maximum value of 20. As a result, only $IF_{\mathrm{dapi}}$ and $IF_{\mathrm{target}}$ can be fully recovered from the inverse transformation.

The offset colors and absorption coefficients are specific to the imaging system and staining protocols used, and were empirically determined based on several factors such as visual quality and similarity to real IHC in-house reference images. **Table S1** shows the values used for different targets.

**Table S1:** Values used for different targets in the pIHC algorithm.

| Value | PanCK | PD-L1 | CD3 | CD8 |
|---|---|---|---|---|
| $B_{\mathrm{r}}$ | 50 | 50 | 50 | 50 |
| $B_{\mathrm{g}}$ | 50 | 50 | 50 | 50 |
| $B_{\mathrm{b}}$ | 50 | 50 | 50 | 50 |
| $W_{\mathrm{r}}$ | 228 | 228 | 228 | 228 |
| $W_{\mathrm{g}}$ | 225 | 225 | 225 | 225 |
| $W_{\mathrm{b}}$ | 233 | 233 | 233 | 233 |
| $\alpha_{\mathrm{dapi,r}}$ | 0.008 | 0.00459 | 0.004 | 0.004 |
| $\alpha_{\mathrm{dapi,g}}$ | 0.006872 | 0.003174 | 0.003436 | 0.003436 |
| $\alpha_{\mathrm{dapi,b}}$ | 0.003376 | 0.001056 | 0.001688 | 0.001688 |
| $\alpha_{\mathrm{target,r}}$ | 0.009 | 0.01119 | 0.009 | 0.009 |
| $\alpha_{\mathrm{target,g}}$ | 0.01992 | 0.02058 | 0.01992 | 0.01992 |
| $\alpha_{\mathrm{target,b}}$ | 0.03 | 0.0294 | 0.03 | 0.03 |
| $\alpha_{\mathrm{residual,r}}$ | 0.0021 | 0.0021 | 0.0021 | 0.0021 |
| $\alpha_{\mathrm{residual,g}}$ | 0.00255 | 0.00255 | 0.00255 | 0.00255 |
| $\alpha_{\mathrm{residual,b}}$ | 0.003 | 0.003 | 0.003 | 0.003 |

#### 2. Hyperparameters

**Table S2** shows the hyperparameter details for the H&E and mIF virtual stainer models.

The learning rate schedule consisted of a linear warmup and cosine decay [44]. Specifically, the learning rate was increased linearly over a certain number of warmup training steps, and decreased at a fixed decay rate with cosine annealing for the remaining training steps.

The loss schedule changed the relative weights of the shift-invariant regression loss and adversarial loss components over time. Specifically, the L1 and L2 regression weights of the shift-invariant regression loss decreased linearly from the initial weight to final weight over the training steps, whereas the conditional and unconditional GAN weights of the adversarial loss increased linearly. If the loss schedule was not employed, the initial and final weights remained the same.

**Table S2:** Hyperparameter values used for the H&E and mIF virtual stainer models.

| Component | Hyperparameter | H&E | mIF |
|---|---|---|---|
| Generator | Kernel size | 3×3 | 3×3 |
| | Number of kernels | 384, 768, 1536, 3072 | 128, 256, 512, 1024 |
| | Dropout | 0.3 | 0.3 |
| | Attention gate | False | True |
| Conditional discriminator | Kernel size | 4×4 | 4×4 |
| | Number of kernels | 384, 768, 1536, 3072 | 128, 256, 512, 1024 |
| | Dropout | 0.1 | 0.1 |
| Unconditional discriminators | Kernel size | 4×4 | 4×4 |
| | Number of kernels | 384, 768, 1536, 3072 | 128, 256, 512, 1024 |
| | Dropout | 0.1 | 0.1 |
| Loss | Schedule | True | False |
| | L1 regression weight (Initial, Final) | 95, 30 | 95, 95 |
| | L2 regression weight (Initial, Final) | 5, 5 | 5, 5 |
| | Conditional GAN weight (Initial, Final) | 0.1, 10 | 100, 100 |

| | | | |
|---|---|---|---|
| | Unconditional GAN1 weight (Initial, Final) | 0.07, 7 | 1, 1 |
| | Unconditional GAN2 weight (Initial, Final) | 0.025, 2.5 | 10, 10 |
| | Rotational consistency weight | 100 | 100 |
| | Regularization weight | 0 | 0.001 |
| Optimizer | Schedule | True | True |
| | Learning rate | 5e-5 | 5e-5 |
| | Warmup steps | 10240 | 10240 |
| | Decay rate | 0.9 | 0.9 |
| | $\boldsymbol{\beta}_1$ | 0.5 | 0.5 |
| | $\boldsymbol{\beta}_2$ | 0.999 | 0.999 |
| Training | Batch size | 16 | 32 |

#### 3. mIF Evaluation Methods

For mIF stains, measurements of the positive cell density, positive cell percentage, TPS, and CPS were calculated based on **Equations S2 - S5**.

$$\text{Positive cell count} = \text{Number of positive cells within region}$$

$$\text{Positive cell density} = \frac{\text{Positive cell count}}{\text{Area of region}} \quad \text{(Equation S2)}$$

$$\text{Positive cell percentage} = 100 \times \frac{\text{Positive cell count}}{\text{Number of cells within region}} \quad \text{(Equation S3)}$$

$$\text{TPS} = 100 \times \frac{\text{Number of PD-L1 positive cells within tumor region}}{\text{Number of cells within tumor region}} \quad \text{(Equation S4)}$$

$$\text{CPS} = 100 \times \frac{\text{Number of PD-L1 positive cells within tissue region}}{\text{Number of cells within tumor region}} \quad \text{(Equation S5)}$$

### 4. mIF Evaluation Results

#### Qualitative Analysis

**Figure S1** shows examples of real and virtual stains of various morphological structures at various magnifications from each individual model. **Figure S2** shows examples of false negatives or false positives in the virtual stains. **Figure S3** shows examples of background and non-specific fluorescence observed in the CD8 real stains which adds label noise during the model training procedure, resulting in poorer performance.

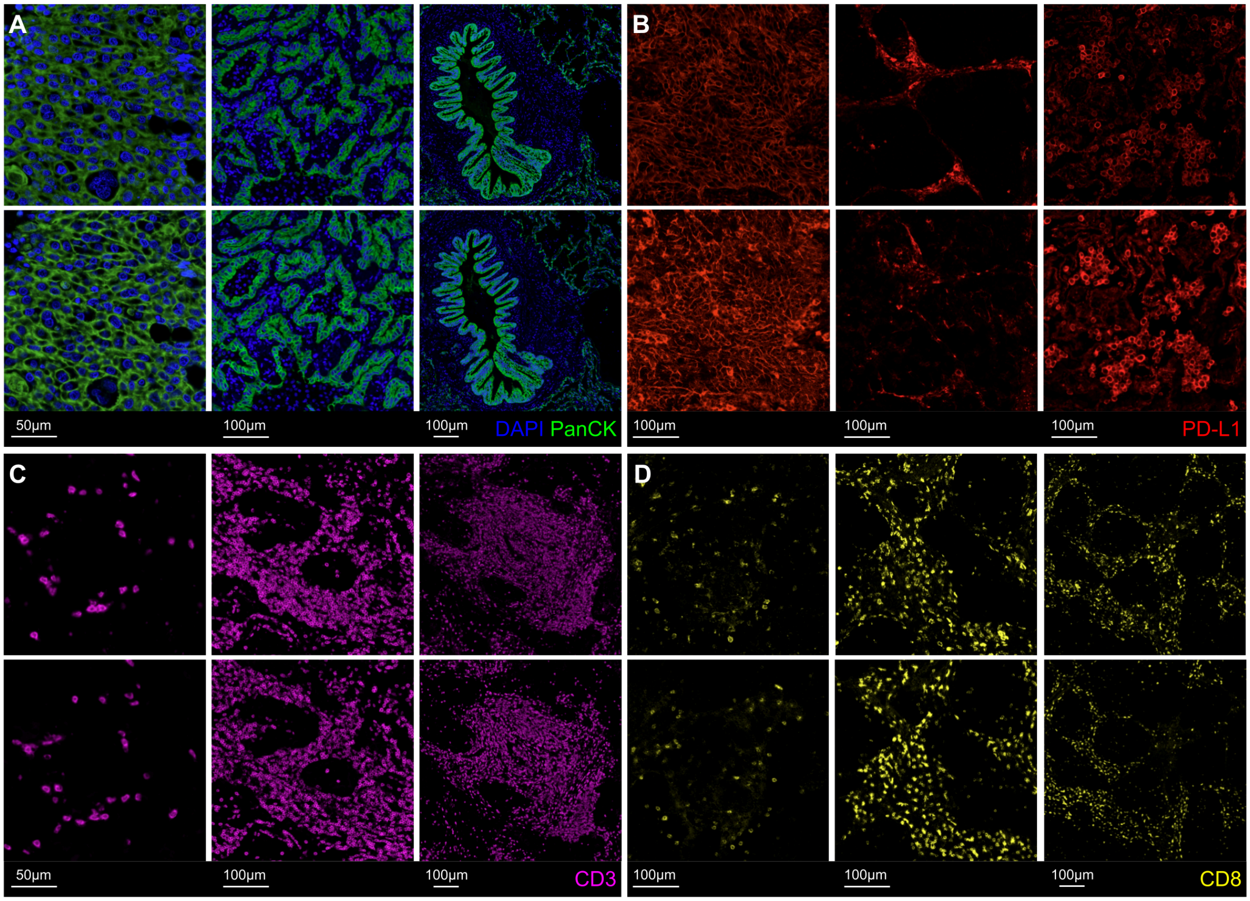


**Figure S1: (A)** Examples of the real (top) and virtual (bottom) stains for DAPI and PanCK showing various morphological features at 40× (left), 20× (middle) and 10× (right) magnifications. **(B)** Examples of the real (top) and virtual (bottom) stains for PD-L1 showing positive expression in tumor cells at 20× magnification (left), negative expression in tumor cells at 20× magnification (middle), and positive expression in immune cells at 20× magnification (right). **(C)** Examples of the real (top) and virtual (bottom) stains for CD3 showing a low density region at 40× magnification (left), high density region at 20× magnification (middle), and tertiary lymphoid structure at 10× magnification (right). **(D)** Examples of the real (top) and virtual (bottom) stains for CD8 showing a low density region at 20× magnification (left), high density region at 20× magnification (middle), and high density region at 10× magnification (right).

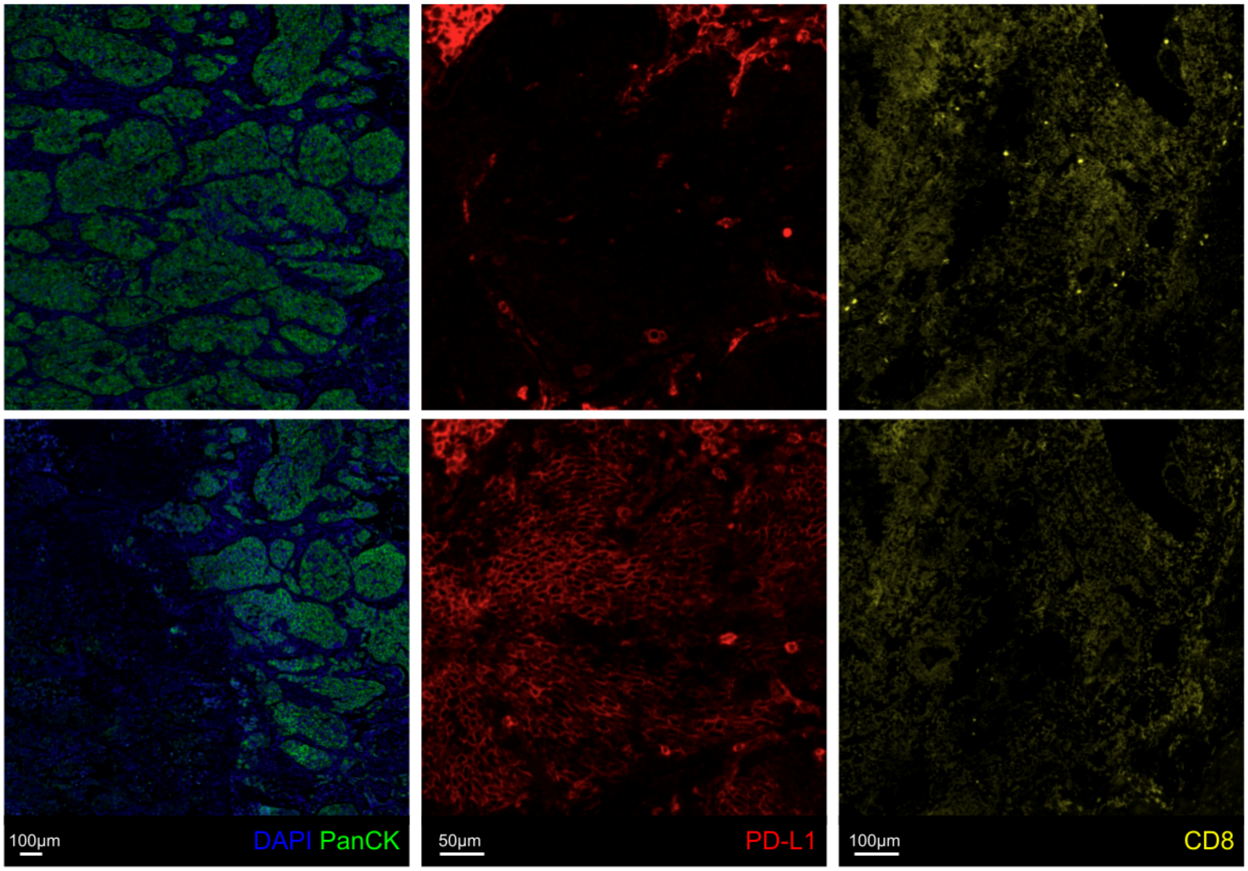


**Figure S2:** Examples of the real (top) and virtual (bottom) stains showing false negative regions for the PanCK virtual stain (left), false positive regions for the PD-L1 virtual stain (middle), and false negative cells for the CD8 virtual stain (right).

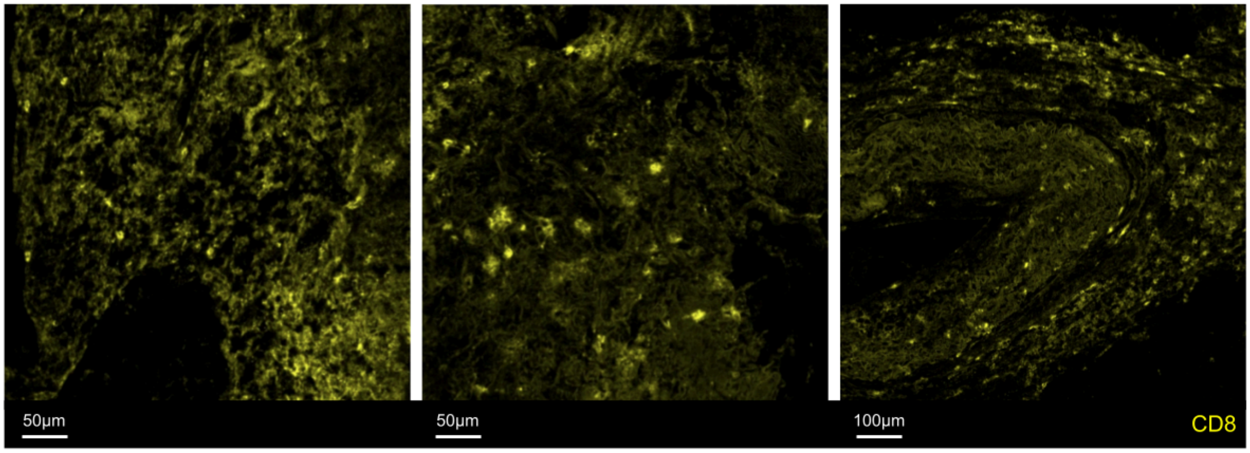


**Figure S3:** Examples of background and non-specific fluorescence observed in CD8 real stains which adds label noise during the model training procedure, resulting in poorer performance.

#### Quantitative Analysis

**Tables S3** and **S4** show the average absolute differences between the measurements on real and virtual stains obtained from the cell segmentation-based analysis in Visiopharm software for the single expression and colocalization analysis, respectively. Blank entries indicate measurements that were not relevant for the stain as described in **Materials and Methods**. **Figures S4 - S11** show the scatterplots of measurements on real and virtual stains.

**Table S3:** Average absolute difference (mean ± SD, median) between measurements on real and virtual stains obtained from the cell segmentation-based analysis in Visiopharm software for PanCK, DAPI, PD-L1, CD3, and CD8 on testing slides. Analysis was performed according to three different definitions of the region of interest.

| Region | Measurement | PanCK | DAPI | PD-L1 | CD3 | CD8 |
|---|---|---|---|---|---|---|
| Tissue | Positive area ($mm^2$) | 6.7 ± 6.5, 4.7 | - | - | - | - |
| | Positive cell density (cells/$mm^2$) | - | 306 ± 388, 201 | 435 ± 357, 340 | 162 ± 170, 111 | 169 ± 188, 104 |
| | Positive cell percentage (%) | - | - | 10.5 ± 6.9, 9.3 | 3.6 ± 3.2, 2.7 | 4.1 ± 4.4, 2.7 |
| Real tumor | Positive cell density (cells/$mm^2$) | - | 557 ± 672, 354 | 820 ± 782, 630 | 205 ± 194, 169 | 180 ± 227, 100 |
| | Positive cell percentage (%) | - | - | See TPS and CPS | 2.9 ± 2.6, 2.2 | 2.8 ± 3.5, 1.7 |
| | TPS (%) | - | - | 11.9 ± 8.6, 11.5 | - | - |
| | CPS (%) | - | - | 18.1 ± 12.1, 16.3 | - | - |
| Respective tumor | Positive cell density (cells/$mm^2$) | - | 845 ± 778, 674 | 834 ± 821, 608 | 278 ± 260, 200 | 192 ± 236, 110 |
| | Positive cell percentage (%) | - | - | See TPS and CPS | 3.6 ± 3.0, 3.0 | 2.8 ± 3.6, 1.7 |
| | TPS (%) | - | - | 11.7 ± 8.7, 11.5 | - | - |
| | CPS (%) | - | - | 20.8 ± 15.5, 16.1 | - | - |

**Table S4:** Average absolute difference (mean ± SD, median) between measurements on real and virtual stains obtained from the colocalization analysis in Visiopharm software for CD3 and CD8, and CD3 and PD-L1, on testing slides. Analysis was performed according to three different definitions of the region of interest.

| Region | Measurement | CD3 and CD8 | CD3 and PD-L1 |
|---|---|---|---|
| Tissue | Positive cell density (cells/mm$^2$) | 94 ± 95, 65 | 112 ± 126, 71 |
| | Positive cell percentage (%) | 2.2 ± 2.1, 1.5 | 2.5 ± 2.3, 1.8 |
| Real tumor | Positive cell density (cells/mm$^2$) | 106 ± 118, 64 | 155 ± 207, 91 |
| | Positive cell percentage (%) | 1.5 ± 1.6, 1.0 | 2.1 ± 2.2, 1.3 |
| Respective tumor | Positive cell density (cells/mm$^2$) | 91 ± 106, 59 | 170 ± 225, 105 |
| | Positive cell percentage (%) | 1.4 ± 1.5, 1.0 | 2.2 ± 2.2, 1.6 |

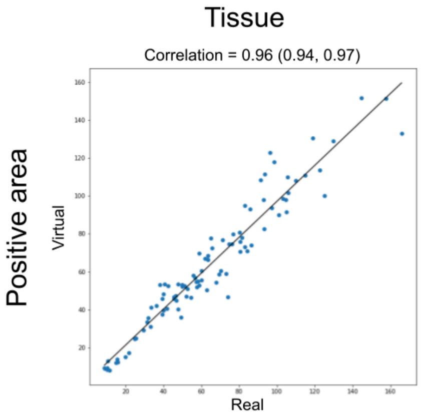


**Figure S4:** Scatterplots of the measurements on real and virtual stains of positive area for PanCK.

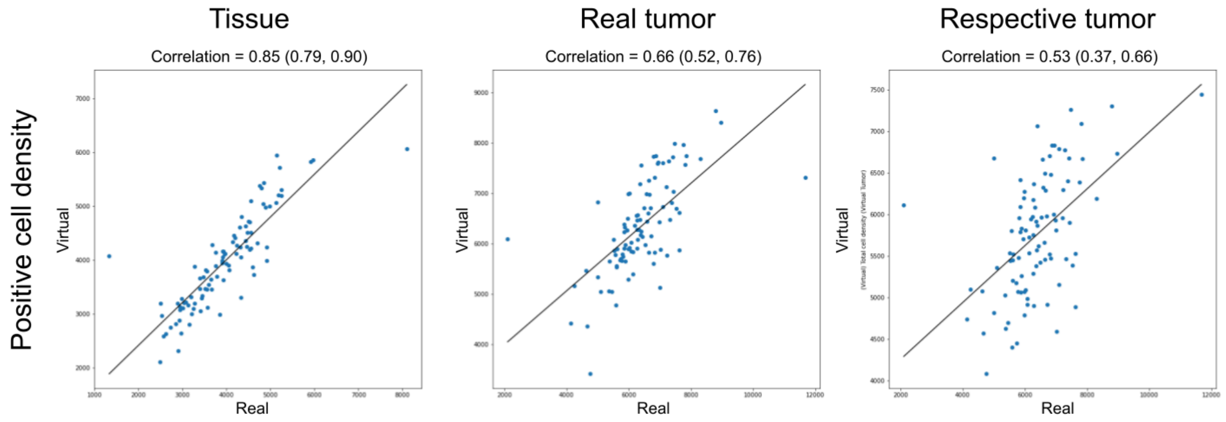


**Figure S5:** Scatterplots of the measurements on real and virtual stains of positive cell density for DAPI.

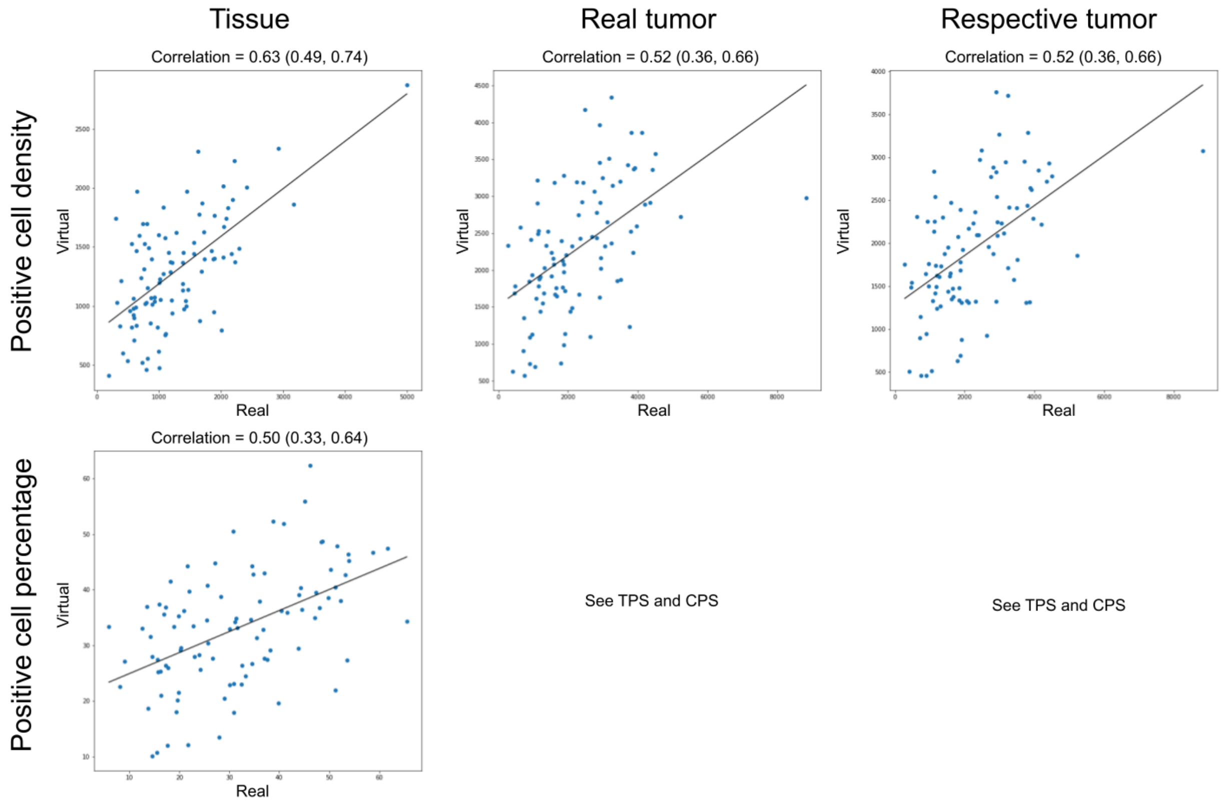


**Figure S6:** Scatterplots of the measurements on real and virtual stains of positive cell density and positive cell percentage for PD-L1.

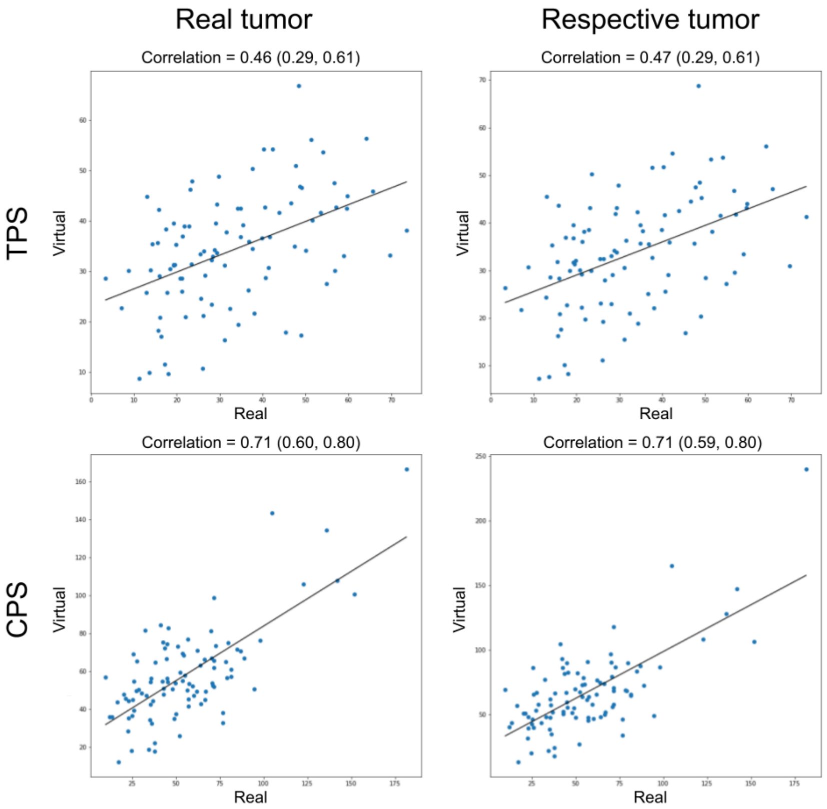


**Figure S7:** Scatterplots of the measurements on real and virtual stains of TPS and CPS for PD-L1.

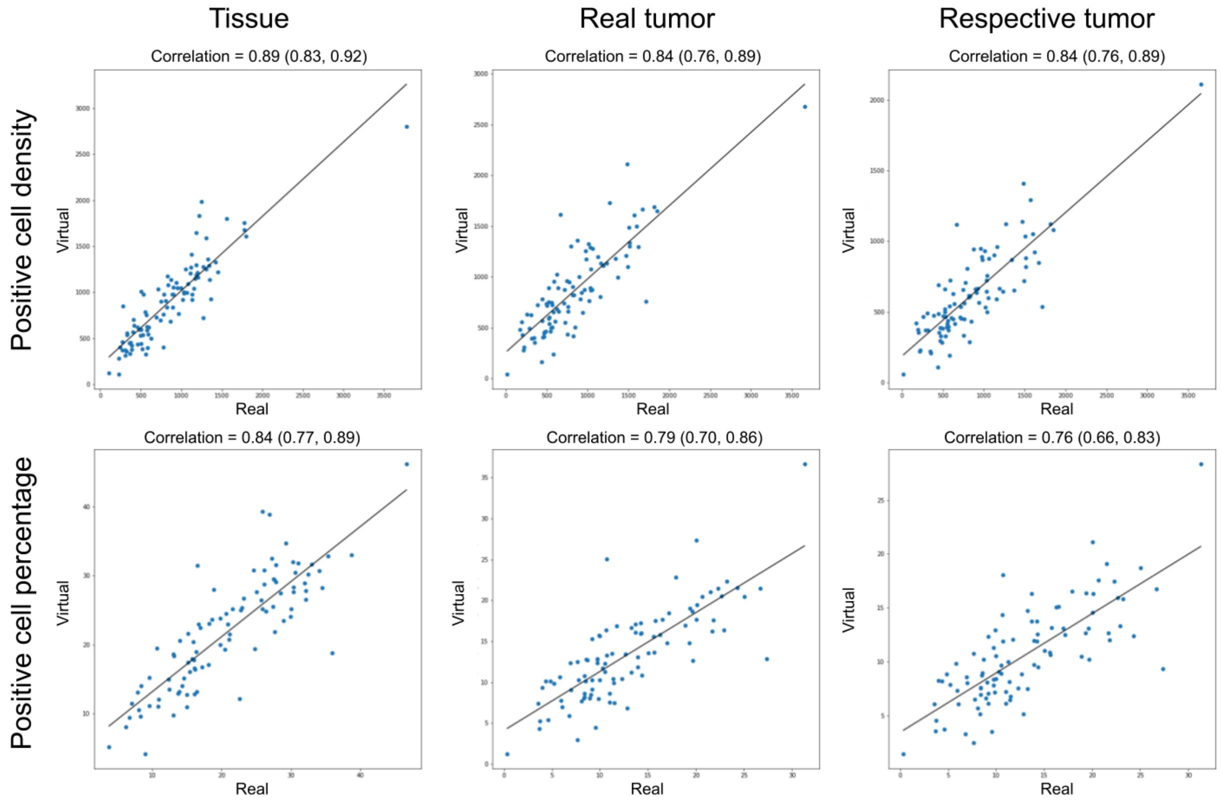


**Figure S8:** Scatterplots of the measurements on real and virtual stains of positive cell density and positive cell percentage for CD3.

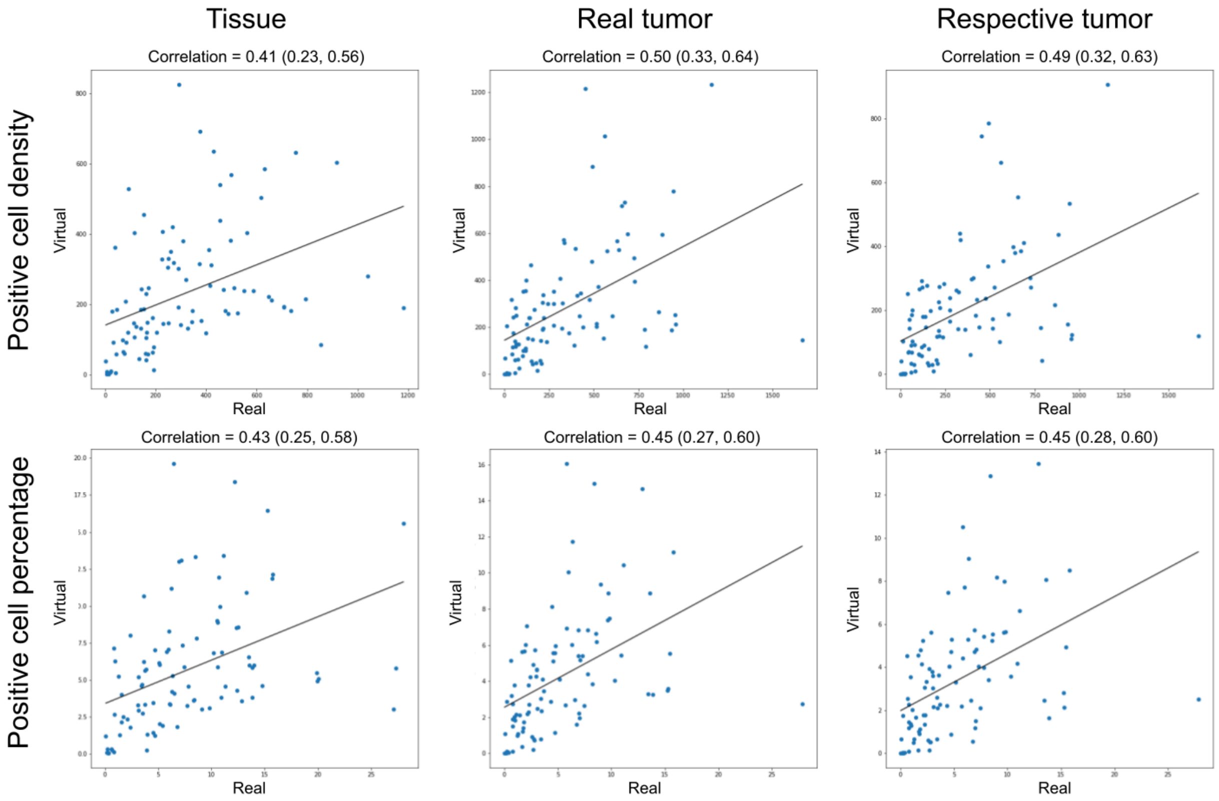


**Figure S9:** Scatterplots of the measurements on real and virtual stains of positive cell density and positive cell percentage for CD8.

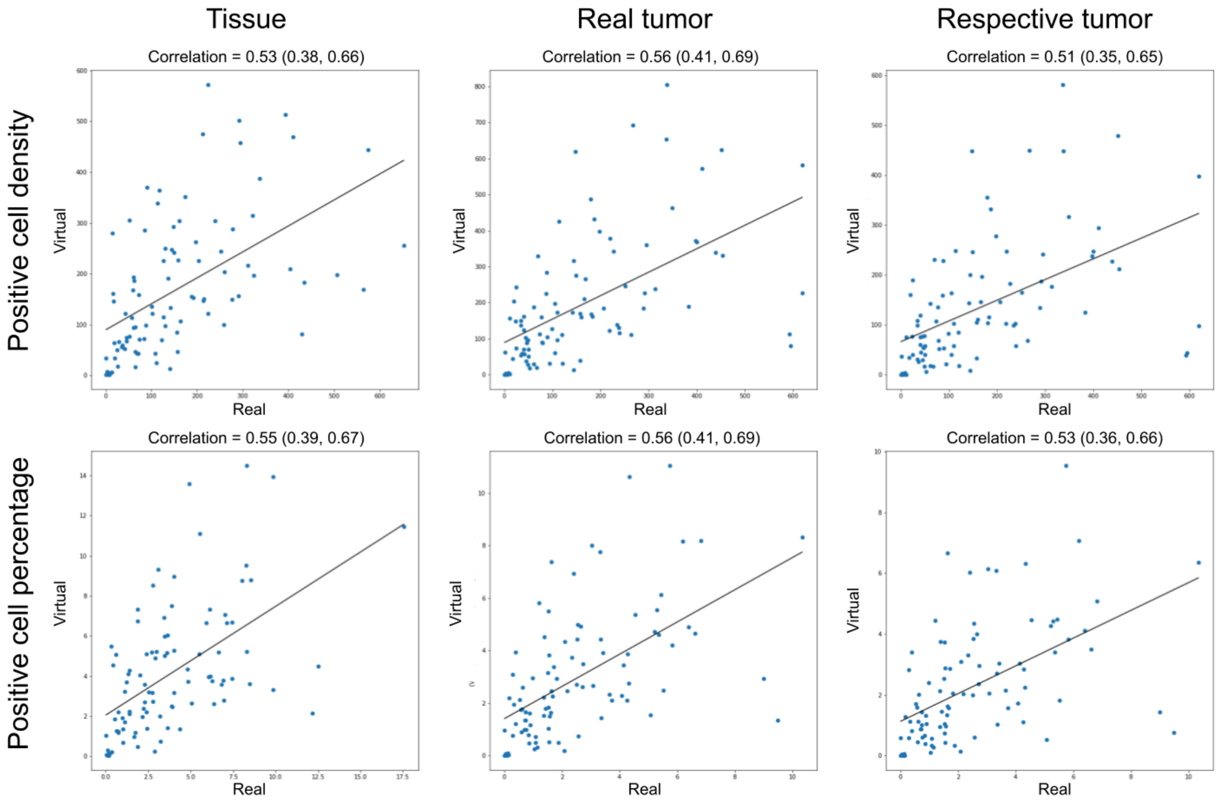


**Figure S10:** Scatterplots of the measurements on real and virtual stains of positive cell density and positive cell percentage for CD3 and CD8 colocalization.

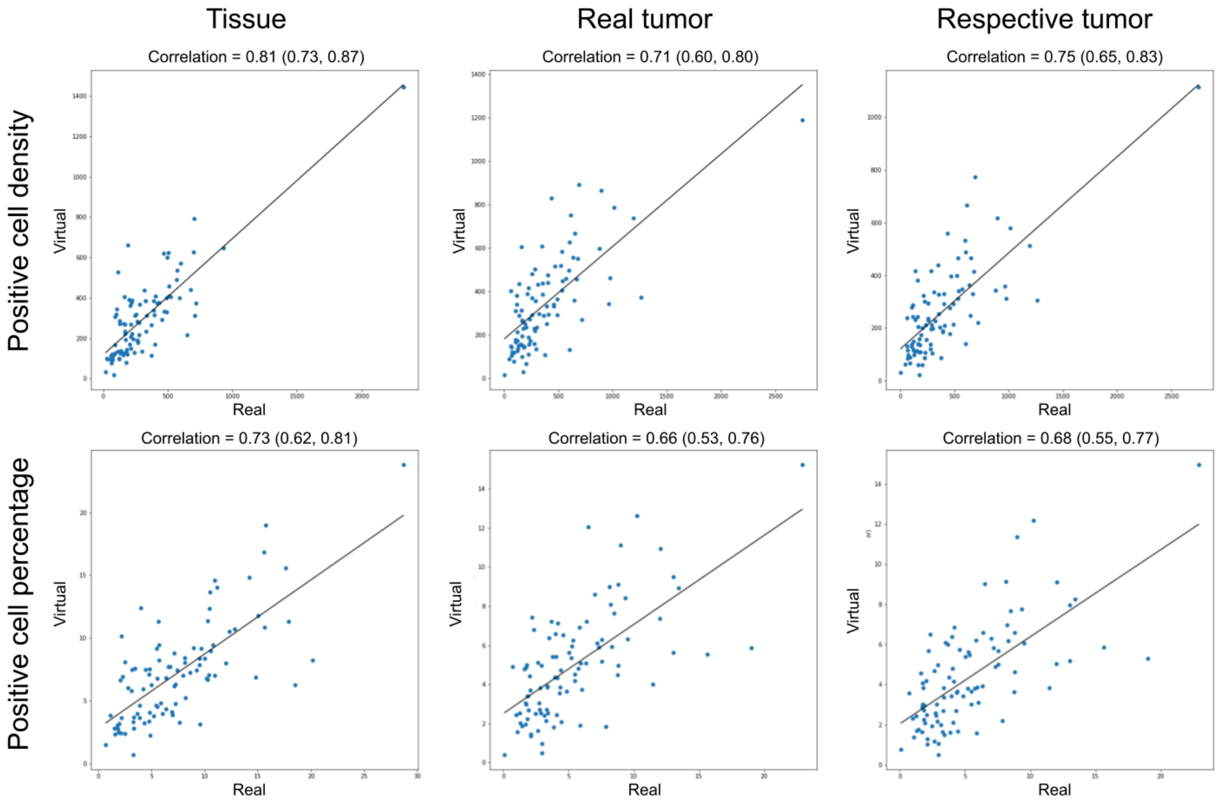


**Figure S11:** Scatterplots of the measurements on real and virtual stains of positive cell density and positive cell percentage for CD3 and PD-L1 colocalization.
